## Appendix 1 for "Knowledge and attitudes about HIV pre-exposure prophylaxis: Evidence from in-depth interviews and focus group discussions with policy makers, healthcare providers, and end-users in Lesotho"

**1. In-depth semi-structured interviews with policy makers**

*As we went over in the consent, all of the information you provide will be kept confidential. Just as a reminder our interview will probably last around 90 minutes. Do you have any questions before we begin?  May I start the recording? [Start recording]*

***Good [afternoon/morning], thank you for participating today****. We are meeting today to talk about your experience with policies around pre-exposure prophylaxis (PrEP) in Lesotho. Remember that your answers are confidential and participation is completely voluntary. Also please keep in mind that there are no right or wrong answers, I am interested in anything you can share with me.*

1. **Opening questions**

a. To begin, I would like to hear more about the work you do, particularly as it relates to HIV/AIDS.
   i. Can you tell me a little more about your position and responsibilities?
   ii. In what ways does PrEP feature in your day-to-day work?
   iii. What other HIV/AIDS-related work do you do?

2. **Introductory questions: attitudes towards PrEP**

a. Today, we will be talking about Pre-Exposure Prophylaxis (PrEP). I would like to get your opinion on PrEP:

i. When did you first hear about it?
ii. What are some of the benefits of the drug?
    1. **Probe:** benefits for individuals; families; community; district; and the country
iii. What are some of the challenges/drawbacks of the drug?
1. **Probe**: challenges for the individuals; families; community; district; and the country
   b. How do you think the general public views PrEP?
i. **Probe**: Positive views; negative views
ii. Why do you think the general public has these views?
iii. How do you think key populations view the drug? [Key populations include Men who have sex with men, Adolescent girls and Young Women, Female sex workers, and other vulnerable populations].

1. Why do you think individuals in these key populations view PrEP this way?

3. **National/district-level current use and vision for PrEP**

a. Could you tell me more about the history of PrEP in Lesotho? And in your district (if applicable)?
   i. When and how was it introduced?
   ii. What were the circumstances that lead to the decision to implement policies around PrEP?
       1. What are some of the pieces of evidence that were used to decide provide PrEP in Lesotho, and in your district (if applicable).
   iii. How would you describe the difference between the facility-based and community-based approaches to providing PrEP?
        1. Probe: what are the advantages and disadvantages of each?
        2. Probe: in your opinion, which has worked better in Lesotho? Please explain why you think that.
b. Currently how would you describe PrEP users in Lesotho? And in your district?
   i. Probe: age range, district, sexual orientation, gender, etc.
   ii. Which populations would you say are being reached for PrEP? Tell me more about that.
   iii. Are there some populations you think should be reached and are not currently?
        1. How or what can we do to reach/inform/educate certain target groups about PrEP?
   iv. What are some reasons you think individuals choose to use PrEP? Tell me more about that.
   v. What are some reasons you think individuals decline the use of PrEP? Tell me more about that.
c. Would you be able to elaborate on the national-level goals of the PrEP program? What about the goals for your district?
   i. **Probe**: Which population are you targeting? Why?
   ii. **Probe**: how may people would you like to reach at a national level? In your district?
   iii. What does it mean for Lesotho to have PrEP available?
   iv. What does it mean for your district to have PrEP available?

4. **PrEP scale up and Implementation**

It is our understanding that the Government of Lesotho, and the Ministry of Health specifically, would like to ensure PrEP is available throughout the Kingdom.

a. Who are the key players/actors in implementing PrEP at the national level? At the district level?
i. **Probe**: What is the actor’s role?
ii. How does your office fit into this PrEP landscape?

**1. Probe:** What role does your office play in implementing PrEP?

b. Could you tell me more about this scale up process? How has the scale up been rolled out?
   i. Probe: criteria for district selection? What are the steps followed to bring PrEP to a community?
      1. What expectations did policy makers have with regard to scaling up PrEP?
   ii. Probe: short-term vision? Long-term vision?
       1. To what extent have those expectations met?
   iii. What would you consider critical elements of the PrEP scale-up?
        1. Probe: health care provider readiness, funding, availability of medication, communication
   iv. To what extent would you say that there is awareness about the PrEP policies at national level? District levels? Among health providers? Among the target populations?
c. How has the scale up experience been so far?
   i. What successes/milestones have you achieved?
   ii. What challenges/barriers have you experienced?
       1. How have you dealt with these challenges?
   iii. What gaps would you say there are in the scale up of PrEP?
        1. How do you think these gaps can be filled?
d. What strategies have been employed to ensure quality is maintained through scale

5. **Lessons learned**

a. Given all that we have talked about today, what are the top 5 pieces of advice you would give to another country planning on providing PrEP to their citizens? Or another district intending to provide PrEP?
i. Probe: things to consider; people to include in discussions/implementation; leadership etc.

6. **Pile sorting exercise**

***Interviewer:*** *use pile sorting and ranking handout to carry out this activity. Record the respondent’s responses in the coversheet.*

7. **Closing**

Thank you for your time. I appreciate your insight on this topic. Before we close, is there any other issue you would like us to discuss?

**2. In-depth semi-structured interviews with implementing partners**

*As we went over in the consent, all of the information you provide will be kept confidential. Just as a reminder our interview will probably last around 90 minutes. Do you have any questions before we begin?  May I start the recording? [Start recording]*

**Good [afternoon/morning], thank you for participating today. We are meeting today to talk about your experience with policies and implementation of pre-exposure prophylaxis (PrEP) in Lesotho. Remember that your answers are confidential and participation is completely voluntary. Also please keep in mind that there are no right or wrong answers, I am interested in anything you can share with me.**

1. **Opening questions**

a. To begin, I would like to hear more about the work you do, particularly as it relates to HIV/AIDS.
   i. Can you tell me a little more about your position and responsibilities?
   ii. In what ways does PrEP feature in your day-to-day work?
   iii. What other HIV/AIDS-related work do you do?

2. **Introductory questions: attitudes towards PrEP and current national/district use**

a. Today, we will be talking about Pre-Exposure Prophylaxis (PrEP). I would like to get your opinion on PrEP:

i. When did you first hear about it?
ii. What are some of the benefits of the drug?
    1. **Probe:** benefits for individuals; families; community; district; and the country
iii. What are some of the challenges/drawbacks of the drug?
     2. **Probe**: challenges for the individuals; families; community; district; and the country
     b. How do you think the general public in Lesotho views PrEP?
i. **Probe**: Positive views; negative views
ii. Why do you think the general public has these views?
iii. How do you think key populations view the drug? [Key populations include Men who have sex with men, Adolescent girls and Young Women, Female sex workers, and other vulnerable populations].

1. Why do you think individuals in these key populations view PrEP this way?

3. **National/district-level vision for PrEP**

a. Could you tell me more about the history of PrEP in Lesotho? And in your district (if applicable)?
   i. When and how was it introduced?
   ii. What were the circumstances that lead to the decision to implement policies around PrEP?
       1. What are some of the pieces of evidence that were used to decide provide PrEP in Lesotho, and in your district (if applicable).
   iii. What successes have you noted to date?
   iv. What challenges have you noted to date?
   v. How would you describe the difference between the facility-based and community-based approaches of providing PrEP?
      1. Probe: what are the advantages and disadvantages of each?
      2. Probe: in your opinion, which has worked better? Please explain why you think that.
b. Currently how would you describe PrEP users?
   i. Probe: age range, district, sexual orientation, gender, etc.
   ii. Which populations would you say are being reached for PrEP? Tell me more about that.
   i. Are there some populations you think should be reached and are not currently?
      1. How or what can we do to reach/inform/educate certain target groups about PrEP?
   ii. What are some reasons you think individuals choose to use PrEP? Tell me more about that.
   iii. What are some reasons you think individuals decline the use of PrEP? Tell me more about that.
b. Would you be able to elaborate on the national-level goals of the PrEP program? What about the goals for your organization?
   i. **Probe**: Which population are you targeting? Why?
   ii. **Probe**: how may people would you like to reach at a national level? In your organization?
   iii. What does it mean for Lesotho to have PrEP available?
   iv. What does it mean for the district you work in to have PrEP available?
c. Who are the key players/actors in implementing PrEP at the national level? At the district level?
   i. **Probe**: What is the actor’s role?
   ii. How does your organization fit into this PrEP landscape?
       1. **Probe:** What role does your office play in implementing PrEP?

4. **PrEP scale up and implementation**

It is our understanding that the Government of Lesotho, and the Ministry of Health specifically, would like to ensure PrEP is available throughout the Kingdom.

a. Could you tell me more about this scale up process? How has the scale up been rolled out?
i. Probe: criteria for district selection?
ii. What expectations did policy makers have with regard to scaling up PrEP?
    1. Probe: short-term vision? Long-term vision?
b. To what extent have those expectations met?
iii. What would you consider critical elements of the PrEP scale-up?
c. Probe: health care provider readiness, funding, availability of medication, communication
iv. To what extent would you say that there is awareness about the PrEP policies at national level? District levels? Among health providers? Among the target populations?
d. How has the scale up experience been so far?
i. What successes/milestones have you achieved?
ii. What challenges/barriers have you experienced?
    1. How have you delt with these challenges?
iii. What gaps would you say there are in the scale up of PrEP?
     1. How do you think these gaps can be filled?
e. What strategies have been employed to ensure quality is maintained through scale

5. **Lessons learned**

a. Given all that we have talked about today, what are the top 5 pieces of advice you would give to another country planning on providing PrEP to their citizens? Or another district intending to provide PrEP?
   a. **Probe**: things to consider; people to include in discussions/implementation; leadership etc.

6. **Pile sorting exercise**

***Interviewer:*** *use pile sorting and ranking handout to carry out this activity. Record the respondent’s responses in the coversheet.*

7. **Closing**

Thank you for your time. I appreciate your insight on this topic. Before we close, is there any other issue you would like us to discuss?

**3. Semi-structured focus group discussions with health providers**

*As we went over in the consent, all of the information you provide will be kept confidential. Just as a reminder our focus group discussion will probably last around 90-120 minutes. Do you have any questions before we begin?  May I start the recording? [Start recording]*

***Good [afternoon/morning], thank you for participating today****. We are meeting today to talk about your experience with pre-exposure prophylaxis (PrEP) in your day-to-day work. Remember that your answers are confidential and participation is completely voluntary. Also please keep in mind that there are no right or wrong answers, I am interested in anything you can share with me.*

1. **Opening questions**

a. To begin, I would like to hear more about the work you do, particularly as it relates to HIV/AIDS. As you mention them, we will list them on the board.
   i. Can you tell me a little more about your positions and responsibilities?
   ii. In what ways does PrEP feature in your day-to-day work?

2. **Introductory questions: attitudes towards, and knowledge of PrEP**

a. Today, we will be talking about Pre-Exposure Prophylaxis (PrEP). I would like to get your opinion on PrEP:

i. What is it? What is it used for? Who is it recommended for?
ii. When did you first hear about it?
iii. What are some of the benefits of the drug?
     1. **Probe:** benefits for individuals; families; community; district; and the country
iv. What are some of the challenges/drawbacks of the drug?
    2. **Probe**: challenges for the individuals; families; community; district; and the country
    b. How do you feel about PrEP as an HIV prevention tool?
    c. To what extent would you say the general public knows about PrEP?
i. How do you think the general public views PrEP?
ii. **Probe**: Positive views; negative views
iii. Does anyone have a story about a time when they overheard a member of the public talk about PrEP?
     1. What did they say?
     2. Why do you think they had these views?

3. **Provider experience, willingness and preparedness to deliver PrEP**

a. What training have you received about PrEP? Please tell me about it.
   i. What did the training entail?
   ii. What would you say was the most beneficial part of the training?
   iii. What do you feel was not as helpful?
   iv. What additional training do you think you still need regarding PrEP?
b. In your health facility, what kinds of conversations occur about PrEP among health providers? Please give examples. How have these conversations changed over time?
   i. To what extent do you think you are capable of providing PrEP in your place of work?
      1. Probe: knowledge of PrEP? Availability of PrEP drug and other supplies? Availability testing facilities? Other?
      2. Probe: who feels they have the tools and skills to provide PrEP? Who feels like they do not? Please tell me more.
   ii. To what extent do you think you are able to manage a client who is on PrEP for a long time?
c. Has any of you present today provided, or counseled a client about PrEP at work? Could you tell me how that experience was? Give examples if possible.
   i. Probe: how did you approach the client? How did you explain how the drug works? How did the client respond? Did the client accept to take the drug? How long was the client on the drug?
   ii. How did the client react when you recommended PrEP?
   iii. Has anyone else had a similar experience?
   iv. Has anyone else had a different experience? Please tell us how your experience differed.
   v. What are some of the things that you have to think about as a provider in this facility whether to offer PrEP?
   vi. What are some of the things that you have to think about as a provider in this facility when to offer PrEP?
   vii. What are some of the things that you have to think about as a provider in this facility how to offer PrEP?
   viii. What are some of the things that you have to think about as a provider in this facility to whom to offer PrEP?
   ix. Has there been a time when you hesitated or decided against offering PrEP?
   x. Has there been a time when you noticed that colleagues hesitated or decided against offering PrEP?
   xi. Has there been a time when you hesitated or decided against offering PrEP?
   xii. Has there been a time when you noticed that colleagues hesitated or decided against offering PrEP?
d. How would you describe the clients you have enrolled on PrEP?
   i. Probe: age range, district, sexual orientation, gender, etc.
   ii. Which populations would you say are being reached for PrEP? Tell me more about that.
   iii. What are some reasons you think individuals choose to use PrEP? Tell me more about that.
e. Could you tell me a story about a time when you counseled a person about PrEP and they declined to enroll.
   i. Please describe the client: e.g. age, socioeconomic status, sex, etc.
   ii. What concerns did the client have about being on PrEP?
   iii. How many of have a similar experience?
   iv. How many have a different experience?
f. How has offering PrEP services affected your day-to-day workload in the facility, if at all?
   i. Is there anything you feel you need (that you currently do not have) that could support you to adequately implement PrEP at your facility?

4. **PrEP adherence and retention**

a. Please think about a client who has done a really good job of adhering to their PrEP medication.
i. Tell me about that client- e.g. age, socioeconomic status, sex, etc.
ii. Why do you think they are able to adhere to their medication?
iii. What are some strategies that they have used to stick to their regiment?
iv. Are there times that they have not adhered? What are the reasons why they missed medication
    i. What are the circumstances that led to them missing a dose?
    ii. What did you do to support the client?
b. Please think about a client who has done a poor job of adhering to their PrEP medication?
   i. Tell me about that client- e.g. age, socioeconomic status, sex, etc.
   ii. Why do you think they are able to adhere to their medication?
   iii. What are some strategies that they have used to stick to their regiment?
   iv. Are there times that they have not adhered? What are the reasons why they missed medication
       i. What are the circumstances that led to them missing a dose?
       ii. What did you do to support the client?

5. **Recommendations**

a. Given all that we have talked about today, and your personal experience dispensing the drug, what are the top 5 pieces of advice you would give to increase the number of clients using PrEP in your facility or community?
i. Probe: what can health providers do? What can health facilities do? What can the government do?
b. If you were sitting with the Ministry and you could tell them just a few things they should think about in terms of challenges you face to offering PrEP, what would be the main things you would want the Ministry to know?

6. **Pile sorting exercise**

***Interviewer:*** *use pile sorting and ranking handout to carry out this activity. Record the respondent’s responses in the coversheet.*

7. **Closing**

Thank you for your time. I appreciate your insight on this topic. Before we close, is there any other issue you would like us to discuss?

**4. In-depth semi-structured interviews with current users of PrEP**

*As we went over in the consent, all of the information you provide will be kept confidential. Just as a reminder our interview will probably last around 90 minutes. Do you have any questions before we begin?  May I start the recording? [Start recording]*

***Good [afternoon/morning], thank you for participating today****. We are meeting today to talk about your experience with using pre-exposure prophylaxis (PrEP). Remember that your answers are confidential and participation is completely voluntary. Also please keep in mind that there are no right or wrong answers, I am interested in anything you can share with me.*

1. **Knowledge, attitudes and decision-making of PrEP**

As I mentioned earlier the focus of our discussion is PrEP. Please think of the first time you heard about PrEP (Pause, await respondent).

a. Please tell me about it – where were you? Who told you about it? Or how did you come to know about it?
   i. What did you think when you heard about this pill?
   ii. Were there any things that you wondered about when you first heard about this pill?
       1. If you could have had more information, what would you have liked to know?
   iii. Please tell me about your first encounter with the health worker who prescribed your PrEP. Describe the interaction.
        1. Probe: what did you think about the counseling you received?
        2. Probe: did you feel like you understood what the health worker was telling you?
        3. Probe: was the counseling clear? What could have made it more clear or better?
        4. Probe: What would you have like to hear about that the health worker did not tell you about?

b. After learning about the pill, how did you decide to start taking it?

i. Probe: Who was involved in the making that decision?

ii. What made you think that PrEP made sense for you?

iii. What made you think that PrEP would be effective?

c. What makes you feel good about being on PrEP?

d. If a friend asked you what PrEP is and how it works, how would you explain it to them? Keep in mind that there are no right or wrong answers

a. **Probe**: Why would a person take PrEP? Why would a person not take PrEP? Who is PrEP for? What else do you know about PrEP?

b. What would be a name that you would give PrEP?
c. When you first started using PrEP, what concerns did you have?

a. **Probe:** to own health? With partner(s)? Larger community? Other?

b. Did those concerns change over the time you have been on PrEP?

d. Do you think PrEP is a topic that men and women in Lesotho feel comfortable discussing with friends and/or partners? Why or why not?

e. Do you know anyone else who is currently taking PrEP? How was their experience starting PrEP different or similar to yours? How did you and this person come to start talking about PrEP?

2. **Preferences in PrEP delivery methods**

a. When you need to collect your pills for PrEP, what will be the steps you need to take? Please take me through the entire process.
i. **Probe:** where do you collect your pills from?
ii. Are there some things that hard to collect the pills?
iii. Are there some things that will make it easy to collect the pills?
b. What do you think the health facility or others who design health promotion programs could do to make it easier for you to collect your PrEP pills?
c. Thinking about Lesotho and your community, what are some things that would make it hard for people like you and people you know to get PrEP? How could things be changed to make it easier for you and others in Swaziland to get and routinely take PrEP?

3. **PrEP adherence and retention**

a. How do you feel about taking this pill every day?
   a. Have there been periods of time when it was difficult for you to take the pill consistently? Could you tell me about those times?
   b. Please remember the last time you missed a dose. Tell me about that day – what was happening? What are the circumstances that led to you missing the dose?
      i. Could you tell me what you did after you missed the dose? Probe: did you take two the next day? Do nothing?
   c. What do you do, with regard to taking the pill, when you are ill from another disease like malaria?
   d. What are some other reasons why it is difficult for you, personally, to be on PrEP?
   e. What are some other reasons why people you know on PrEP find it difficult to take the drug?
b. What are some strategies have you used to remember to take the pill everyday?
   a. Probe: if your fiend was having trouble remembering to take the pill every day, what advise would you give them?
c. What do you think will make it hard or easy for you (and/or others you know) to take the pill as often as the provider recommends?
   a. What do you think would be useful to improve this for you or others?
d. What could people who oversee health interventions do to support you and others in taking PrEP?

4. **Effect of PrEP on life**

Now, I would like to discuss something a bit more intimate. Please remember that all information you share is confidential and will only be shared with the research team for purposes of improving access to PrEP. Ok? (pause). I would like to talk about your intimate life in relation to PrEP (pause).

a. How has PrEP affected your sexual life?
   a. **Probe**: Has it affected or might it affect, whether or how you discuss HIV status with partners?
   b. **Probe**: How might it effect whether you use condoms?
   c. **Probe**: How might it effect on the number of sexual partners you may have?
b. Is there a time earlier in life when, looking back now, you wish you would have had access to PrEP? Please tell me about it.
c. Would you recommend PrEP to your closest friend? Please explain why? Or why not?

5. **Pile sorting exercise**

***Interviewer:*** *use pile sorting and ranking handout to carry out this activity. Record the respondent’s responses in the coversheet.*

6. **Closing**

a. To conclude, what would be your recommendation to improve your experience with PrEP?
b. What would you recommend to improve others’ experience and access to PrEP?

Thank you for your time. I appreciate your insight on this topic. Before we close, is there any other issue you would like us to discuss?

**5. In-depth semi-structured interviews with former PrEP users**

*As we went over in the consent, all of the information you provide will be kept confidential. Just as a reminder our interview will probably last around 90 minutes. Do you have any questions before we begin?  May I start the recording? [Start recording]*

***Good [afternoon/morning], thank you for participating today****. We are meeting today to talk about your experience with using pre-exposure prophylaxis (PrEP). Remember that your answers are confidential and participation is completely voluntary. Also please keep in mind that there are no right or wrong answers, I am interested in anything you can share with me.*

1. **Knowledge, attitudes and decision-making of PrEP**

As I mentioned earlier the focus of our discussion is PrEP. Please think of the first time you heard about PrEP (Pause, await respondent).

a. Please tell me about it – where were you? Who told you about it? Or how did you come to know about it?
   i. What did you think when you heard about this pill?
   ii. Were there any things that you wondered about when you first heard about this pill?
       1. If you could have had more information, what would you have liked to know?

b. If a friend asked you what PrEP is and how it works, how would you explain it to them? Keep in mind that there are no right or wrong answers

i. **Probe**: Why would a person take PrEP? Why would a person not take PrEP? Who is PrEP for? What else do you know about PrEP?
c. What would be a name that you would give PrEP?
d. Do you think PrEP is a topic that men and women in Lesotho feel comfortable discussing with friends and/or partners? Why or why not?

e. After learning about the pill, how did you decide to start taking it?

i. Probe: Who was involved in the making that decision?

ii. Probe: What made you think that PrEP made sense for you?

iii. Probe: What made you think that PrEP would be effective?

f. When you first started using PrEP, what concerns did you have?

i. **Probe:** to own health? With partner(s)? Larger community? Other?

ii. Did those concerns change over the time you have been on PrEP?

g. Please tell me about your first encounter with the health worker who prescribed your PrEP. Describe the interaction.
   i. Probe: what did you think about the counseling you received?
   ii. Probe: did you feel like you understood what the health worker was telling you?
   iii. Probe: was the counseling clear? What could have made it more clear or better?
   iv. Probe: What would you have like to hear about that the health worker did not tell you about?

h. Could you please take me through your decision to stop taking PrEP. What are the circumstanced that led to that decision?

i. Probe: Who was involved in the making that decision?

ii. Probe: what are the reasons why to decide to stop taking PrEP? access? Perceived risk? Change in relationship status?

iii. How settled are you in your decision to stop taking PrEP? Do you think you would ever go back to taking it? Why? Why not?

i. Overall, how would you describe your experience with PrEP for the time you used it?

i. Do you know anyone else who is currently taking PrEP? How was their experience using PrEP different or similar to yours? How did you and this person come to start talking about PrEP?

2. **Preferences in PrEP delivery methods**

a. When you needed to collect your pills in the past for PrEP, what will be the steps you need to take? Please take me through the entire process.
i. **Probe:** where did you collect your pills from?
ii. Are there some things that hard to collect the pills?
iii. Are there some things that will make it easy to collect the pills?
b. What do you think the health facility or others who design health promotion programs could do to make it easier for those interested in PrEP to collect it?
c. Thinking about Lesotho and your community, what are some things that would make it hard for people like you and people you know to get PrEP? How could things be changed to make it easier for you and others in Swaziland to get and routinely take PrEP?

3. **PrEP adherence and retention**

a. How did you feel about taking this pill every day?
   a. Were there periods of time when it was difficult for you to take the pill consistently? Could you tell me about those times?
   b. Please remember the last time you missed a dose. Tell me about that day – what was happening? What are the circumstances that led to you missing the dose?
      i. Could you tell me what you did after you missed the dose? Probe: did you take two the next day? Do nothing?
   c. What do you do, with regard to taking the pill, when you are ill from another disease like malaria?
   d. What are some other reasons why it is difficult for you, personally, to be on PrEP?
   e. What are some other reasons why people you know on PrEP find it difficult to take the drug?
b. What are some strategies you used to remember to take the pill everyday?
   a. Probe: if your fiend was having trouble remembering to take the pill every day, what advise would you give them?
c. What do you think will make it hard or easy for you (and/or others you know) to take the pill as often as the provider recommends?
   a. What do you think would be useful to improve this for you or others?
d. What could people who oversee health interventions do to support you and others in taking PrEP?

4. **Effect of PrEP on life**

Now, I would like to discuss something a bit more intimate. Please remember that all information you share is confidential and will only be shared with the research team for purposes of improving access to PrEP. Ok? (pause). I would like to talk about your intimate life in relation to PrEP (pause).

a. How did PrEP affected your sexual life?
   a. **Probe**: Has it affected or might it affect, whether or how you discuss HIV status with partners?
   b. **Probe**: How might it effect whether you use condoms?
   c. **Probe**: How might it effect on the number of sexual partners you may have?
b. Is there a time earlier in life when, looking back now, you wish you would have had access to PrEP? Please tell me about it.
c. Would you recommend PrEP to your closest friend? Please explain why? Or why not?

5. **Pile sorting exercise**

***Interviewer:*** *use pile sorting and ranking handout to carry out this activity. Record the respondent’s responses in the coversheet.*

6. **Closing**

a. To conclude, what would be your recommendation to improve your experience with PrEP?
b. What would you recommend to improve others’ experience and access to PrEP?

Thank you for your time. I appreciate your insight on this topic. Before we close, is there any other issue you would like us to discuss?

**6. In-depth semi-structured interviews with decliners of PrEP**

*As we went over in the consent, all of the information you provide will be kept confidential. Just as a reminder our interview will probably last around 90 minutes. Do you have any questions before we begin?  May I start the recording? [Start recording]*

***Good [afternoon/morning], thank you for participating today****. We are meeting today to talk about your experience with using pre-exposure prophylaxis (PrEP). Remember that your answers are confidential and participation is completely voluntary. Also please keep in mind that there are no right or wrong answers, I am interested in anything you can share with me.*

1. **Opening questions**

As I mentioned earlier the focus of our discussion is PrEP. Please think of the first time you heard about PrEP.

a. Do you remember the first time you heard about it? (Pause, await respondent). Please tell me about it.
b. Keeping in mind that there are no right or wrong answers, in your understanding, what is PrEP?
c. What would be a name that you would give PrEP?

d. If a friend asked you how PrEP works, how would you explain it to them?

i. Probe: Why would a person take PrEP?
ii. Probe: Why would a person not take PrEP?
iii. Probe: Who is PrEP for?
e. What else do you know about PrEP?

2. **Knowledge, attitudes and decision-making of PrEP**

Now please walk me through your story from when you heard about PrEP until now. If you don’t mind, I’ll interrupt sometimes to get more details.

a. When and where did you first hear about PrEP?

i. What did you think when you heard about this pill?
ii. Were there any things that you wondered about when you first heard about this pill?
iii. If you could have had more information, what would you have liked to know?

iv. What made you think that PrEP did not make sense for you?

v. What made you think that PrEP would not be effective?

b. Could you please tell be about the day you were offered PrEP?

i. Probe: tell me about the day? Where were you? Who talked to you? What did they say?

ii. How did you make your decision not to accept PrEP?

1. Probe: who was involved in making the decision?

c. Please tell me about your first encounter with the health worker who prescribed your PrEP. Describe the interaction.
   i. Probe: what did you think about the counseling you received?
   ii. Probe: did you feel like you understood what the health worker was telling you?
   iii. Probe: was the counseling clear? What could have made it more clear or better?
   iv. Probe: What would you have like to hear about that the health worker did not tell you about?

d. Do you think PrEP is a topic that men and women in Lesotho feel comfortable discussing with friends and/or partners? Why or why not?

e. Do you know anyone else who is currently taking PrEP? How was their experience starting on PrEP different or similar to yours? How did you and this person come to start talking about PrEP?

a. Let us imagine that there is a woman. She does not know her husband’s HIV status. She thinks he may have other sexual partners.
   a. Would it make sense for her to take PrEP?
   b. What would be some of the benefits for a person like her taking PrEP?
   c. What would be some of the drawbacks?

a. Let us imagine that there is a man. He knows his wife is HIV positive. He is negative.
   a. Would it make sense for him to take PrEP?
   b. What would be some of the benefits for a person like him taking PrEP?
   c. What would be some of the drawbacks of him taking PrEP?

3. **Reasons for decline**

a. When you first started using PrEP, what concerns did you have?

a. **Probe:** to own health? With partner(s)? Larger community? Other?

b. Did those concerns change over the time you have been on PrEP?

b. Were there any things that made you worried or concerned about starting PrEP?

There are a lot of reasons that people take or don’t take medicine even if providers would like people to take a medicine. The Ministry of Health wants to learn how to make more Basotho interested in taking PrEP. The people in the Ministry can’t talk to everyone in Lesotho, so the thoughts you can share are very valuable to me. Your opinion represents many many people who we cannot talk to directly.

a. In your opinion, why do you think some people – even those who are at risk for HIV – do not want to take PrEP?
b. How could the Ministry get more people interested in PrEP?
c. How could things be changed to make it easier for you and others in Lesotho to get and routinely take PrEP?
d. Can you imagine a time in the coming months or years when you might be willing to come back to learn more about PrEP or to request PrEP medicines?

4. **PrEP adherence and retention**

a. How do you feel about taking a pill every day?
   i. Can you think of periods of time when it was difficult for you to take the pill consistently?
   ii. Could you imagine what you would do if you missed a dose?
   iii. What would you do, with regard to taking the pill, when you are ill from another disease like malaria?
   iv. What are some other reasons why people you know on PrEP find it difficult to take the drug?
b. What are some strategies you think could be used to remember to take the pill everyday?
   i. Probe: if your fiend was having trouble remembering to take the pill every day, what advise would you give them?
c. What do you think will make it hard or easy for someone (and/or others you know) to take the pill as often as the provider recommends?
   i. What do you think would be useful to improve this for you or others?
d. What could people who oversee health interventions do to support you and others in taking PrEP?

5. **Pile sorting exercise**

***Interviewer:*** *use pile sorting and ranking handout to carry out this activity. Record the respondent’s responses in the coversheet.*

6. **Closing**

a. To conclude, what would be your recommendation to improve your experience with PrEP?
b. What would you recommend to improve others’ experience and access to PrEP?

Thank you for your time. I appreciate your insight on this topic. Before we close, is there any other issue you would like us to discuss?
