## Appendix 2 for "Knowledge and attitudes about HIV pre-exposure prophylaxis: Evidence from in-depth interviews and focus group discussions with policy makers, healthcare providers, and end-users in Lesotho"

**Appendix 2:** Qualitative Codebook

| **Topic/Theme** | **Code** | **Description** |
| --- | --- | --- |
| **Participant characteristics** | Policy maker | Applied to the entire transcript to identify the respondent as a policy maker |
|  | Implementing partner | Applied to the entire transcript to identify the respondent as an implementing partner |
|  | Health provider | Applied to the entire transcript to indicate that it was a focus group discussion with health providers |
|  | Current user | Applied to the entire transcript to identify the respondent as a current PrEP user |
|  | Former user | Applied to the entire transcript to identify the respondent as a former PrEP user |
|  | Decliner | Applied to the entire transcript to identify the respondent as a PrEP decliner |
|  | Urban | Applied to the entire transcript to indicate that this interview or focus group discussion was conducted in an urban location |
|  | Rural | Applied to the entire transcript to indicate that this interview or focus group discussion was conducted in a rural location |
|  | Female | Applied to the entire transcript to indicate that this interview was conducted with a respondent who identifies as female |
|  | Male | Applied to the entire transcript to indicate that this interview was conducted with a respondent who identifies as male |
| **Quote** | Quote | Piece of intriguing data depicting relevant theme that could potentially be quoted in a manuscript |
| **Attitudes towards PrEP** | Attitudes_negative | Personal or otherwise encountered negative perceptions of PrEP expressed by the respondent |
|  | Attitudes_neutral | Personal or otherwise encountered neutral perceptions of PrEP expressed by the respondent |
|  | Attitudes_positive | Personal or otherwise encountered positive perceptions of PrEP expressed by the respondent |
|  | Attitudes_name | A Sesotho name or phrase respondent provided to describe or encapsulate PrEP as an idea and/or drug |
|  | Attitudes_other | Any other data of perceptions and attitudes expressed by the respondent not captured in other codes |
| **PrEP knowledge** | Knowledge_understanding | Knowledge, understanding and/or perception of how PrEP works in the body to prevent HIV infection |
|  | Knowledge_user | Knowledge PrEP users have gained through media messaging the receipt of counseling or other avenues |
|  | Knowledge_other | Any other data related to PrEP knowledge not captured in other codes |
| **PrEP misconceptions** | Misconception_HIV+ | The misconception, either personally held or otherwise encountered, that PrEP leads to the individual taking the drug to be HIV positive |
|  | Misconception_once | The misconception, either personally held or otherwise encountered, that one dose of PrEP offers life-long protection |
|  | Misconception_pregnancy | The misconception, either personally held or otherwise encountered, that PrEP can protect against pregnancy |
|  | Misconceptions_STIs | The misconception, either personally held or otherwise encountered, that PrEP can protect against STIs |
|  | Misconception_other | Any data related misconceptions around PrEP not captured in other codes |
